# Long-term mortality trends among individuals with tuberculosis: a retrospective cohort study of individuals diagnosed with tuberculosis in Brazil

**DOI:** 10.1101/2024.11.20.24317659

**Authors:** Sun Kim, Daniele M. Pelissari, Luiza O. Harada, Mauro Sanchez, Patricia Bartholomay Oliveira, Fernanda D. C. Johansen, Ethel L. N. Maciel, Ted Cohen, Marcia C. Castro, Nicolas A. Menzies

## Abstract

**Background:** Even after successful treatment, tuberculosis (TB) survivors experience elevated morbidity and mortality. However, there is limited evidence on how these risks vary over time and according to individual characteristics.

**Methods:** We conducted a retrospective cohort study of individuals diagnosed with TB in Brazil, using national TB notifications and linked mortality records for 2007-2016. For this study population we estimated mortality rate ratios (MRRs) and cumulative mortality by year since TB diagnosis, as compared to general population mortality rates matched on age, sex, year, and state. We identified clinical and sociodemographic factors associated with elevated post-TB mortality, and compared the distribution of causes of death to the general population.

**Findings:** The study sample included 834,594 individuals, with 4.1 million person-years of follow-up. The TB cohort had elevated mortality compared to the general population, particularly in the first year post-diagnosis (MRR 11.28, 95%CI: 11.18–11.37). Post-TB MRRs declined from 3.59 (3.53–3.64) in year 2 to 1.46 (1.34–1.59) in year 10. Cumulative excess mortality was 6.12% (95%CI: 6.07–6.17) after 1 year and 9.90% (9.58–10.24) after 10 years. MRRs were highest for individuals 30-44 years-old at diagnosis. Relapse, loss to follow-up, and co-prevalent conditions like HIV and alcoholism were strongly associated with higher MRRs. Over time, causes of death shifted from TB and HIV to chronic conditions like cardiovascular disease and cancer.

**Interpretation:** Individuals developing TB disease face elevated mortality up to 10 years after diagnosis. These excess risks vary across demographic and clinical characteristics.

**Funding:** NIH.

## INTRODUCTION

Tuberculosis (TB) remains one of the leading causes of death from a single infectious agent globally, responsible for 1.25 million deaths in 2023.^1^ Although effective TB treatment can reduce TB-related deaths, many individuals who survive TB disease experience elevated morbidity and mortality many years after TB cure. Observational studies have consistently estimated that TB survivors have higher all-cause mortality rates compared to the general population.^2–6^ This elevated mortality results from chronic sequelae caused by TB disease,^7–9^ as well as the effects of pre-existing differences in health status, health behaviors, and social conditions between individuals who develop TB and the general population.^10^ However, there is limited evidence describing how the excess mortality faced by TB survivors varies over time since TB or according to individual characteristics. Given the substantial number of individuals living with a history of TB disease^11^ and the significant health losses in this population,^12^ understanding how long-term mortality trends among TB survivors vary across the cohort could allow better quantification of future health risks and identify clinical and patient factors that could be targeted to improve long-term outcomes.

In this study we estimated mortality trends among individuals with TB, from the point of TB diagnosis, during TB treatment, and after treatment completion. To do so we used data from Brazil’s national TB notification system linked to mortality records from 2007 to 2016. For this TB cohort we estimate differences in mortality rates over time since TB diagnosis as compared to the general population, and analyzed how mortality trends differed by age group, sex, and other sociodemographic and clinical factors. We calculated differences in cumulative mortality to assess how deaths during the TB episode and deaths during the post-TB period contributed to overall excess mortality, and described how causes of death changed over time.

## METHODS

### Linked TB diagnosis and mortality data

We utilized a linked dataset of tuberculosis (TB) case notifications from Brazil’s National Notifiable Disease Information System *SINAN* (*Sistema de Informação de Agravos de Notificação*)^13^ for the period 2007-2016, and death records for any individual with notified TB included in the SINAN TB dataset, as recorded in the Brazilian Mortality Information System *SIM* (*Sistema de Informação sobre Mortalidade*),^14^ covering the same period. The linkage of disease diagnosis and mortality data was conducted by surveillance staff at the Brazilian Ministry of Health.^15^ For each individual with diagnosed TB, the linked SINAN–SIM dataset includes socio-demographic and clinical information recorded at the time of TB diagnosis and treatment. We extracted variables for date of diagnosis, age at diagnosis, sex, self-reported race, state of residence, educational attainment, presence of co-prevalent medical and social conditions (HIV, alcoholism, diabetes, mental illness, and incarceration), and features of diagnosis and treatment (diagnostic test results, type of TB (pulmonary/extrapulmonary), chest X-ray result, and recorded treatment outcome (cured, loss to follow-up, transfer, treatment failure, drug resistance, other). The dataset also included variables from death records, if a death was recorded for the individual over the 2007-2016 period. For individuals with a recorded death, we extracted the date of death and cause of death.

We excluded individuals whose TB diagnosis was subsequently revised to a different disease diagnosis (these individuals were assumed to not have TB) and individuals who received a diagnosis of TB postmortem. Additionally, we excluded records with missing age, sex, or state, and individuals aged 90 years or older.

Individuals were censored at the point of death, and were otherwise assumed to remain in the study cohort. This approach assumed (a) that individuals do not emigrate (since deaths occurring outside Brazil would not be recorded in SIM), and (b) that all deaths in Brazil are recorded in SIM with sufficient detail to be accurately linked to a SINAN record.

### General population data

To estimate mortality rates for the general population we extracted reported death counts from *SIM* for the years 2007 to 2016 along with population data for the corresponding year, age, sex, and state from the Brazilian Institution of Geography and Statistics *IBGE* (*Instituto Brasileiro de Geografia e Estatística*).^16^ Using these data we fitted generalized additive regression models to create smoothed mortality estimates for the general population stratified by year, age, sex, and state. Mortality rates in the TB cohort were compared to these general population mortality rates to estimate the additional mortality faced by individuals diagnosed with TB.

### Outcomes of interest

The primary outcome was mortality rate ratios (MRRs) for individuals in the TB cohort compared to the general population, for each year since TB diagnosis. We estimated this outcome for the overall cohort as well as by age groups based on age at TB diagnosis (ages 0-14, 15-29, 30-44, 45-59, 50-74, and 75-89 years), by sex (female and male), and by region (North, Northeast, Central-West, Southeast, and South). We defined the ‘TB episode’ as the 12 months following TB diagnosis, and the ‘post-TB period’ as any observation time >12 months following diagnosis.^6^ We fit univariable and multivariable regression models for mortality during the TB episode and during the post-TB period, respectively, to estimate the relationship between MRRs and a range of socio-demographic and clinical factors, including sex, bacteriological test results, type of TB (pulmonary vs. extrapulmonary), chest X-ray result, type of entry (new case or relapse), comorbid conditions, diagnosis in prison, and recorded treatment outcome (cured, loss to follow-up, treatment failure, transfer, change in treatment, and unknown).

Using the estimated MRRs and general population mortality rates, we estimated cumulative mortality over time for the TB cohort compared to the general population. We calculated the difference between these cumulative mortality curves to quantify the excess mortality experienced by the TB cohort over the follow-up period. To quantify the relative contribution of deaths during the TB episode and deaths during the post-TB period to overall excess mortality, we estimated excess mortality under an additional counterfactual scenario that only applied TB-associated MRRs over the TB period (first 12 months after the date of TB diagnosis).

In addition, we analyzed the cause of death distribution using ICD-10 codes, categorizing deaths into 11 groups (TB, HIV, respiratory disease, trauma, endocrine/metabolic disorder, digestive disease, cardiovascular disease, cancer, other, and unknown). For the TB cohort, the proportion of deaths by cause was calculated for each year since TB diagnosis. We compared these results to the distribution of deaths in the general population matched by age, sex, state, and calendar year.

### Statistical analysis

Study outcomes were estimated from generalized linear regression models fit with a complementary log-log link function and an offset term calculated as the log of the expected number of deaths in the general population. In this regression framework, the exponentiated regression coefficients represent MRRs for the TB cohort compared to the general population. We estimated equal-tailed 95% confidence intervals by simulating from the uncertainty distributions of the model coefficients. All analyses were conducted in R v4.4.1,^17^ using the mgcv package v1.9-1.^18^

## RESULTS

Between 2007 and 2016 a total of 861,928 tuberculosis cases were recorded in the SINAN database. After excluding cases with unknown age (n=344), age 90 or older (n=1,725), missing sex (n=53), missing state of residence (n=324), postmortem diagnosis (n=6,041), or a recorded change in diagnosis (n=18,847), the final dataset included 834,594 individuals with TB. The average duration of follow-up was 1,789 days (4.9 years), resulting in a total of 4,089,883 person-years of follow-up. There were 120,330 deaths recorded in the study cohort over this follow-up period (14% of individuals in the cohort). **Table 1** summarizes the characteristics of individuals included in the study cohort, including the number of deaths and person-years of follow-up during the observation period. In addition, **Table S1** reports the prevalence of comorbid conditions (HIV, diabetes, alcoholism, and mental illness) across age groups in the TB cohort.

**Table 1.** Characteristics of the cohort.

|  | Number of individuals<br>in cohort | All-cause deaths (%)<br>at the end of the follow-up | Number of person-years of follow-up |
| --- | --- | --- | --- |
| **Total** | **834,594** | **120,330 (14.4%)** | **4,089,883** |
| **Year of diagnosis** |  |  |  |
| 2007-2010 | 263,363 | 61,033 (23.2%) | 2,426,476 |
| 2011-2013 | 221,066 | 36,919 (16.7%) | 1,180,657 |
| 2014-2016 | 229,835 | 22,378 (9.7%) | 482,750 |
| **Age group** |  |  |  |
| 0-14 | 25,077 | 1,014 (4.0%) | 143,924 |
| 15-29 | 228,803 | 15,187 (6.6%) | 1,275,255 |
| 30-44 | 229,154 | 34,537 (15.1%) | 1,305,020 |
| 45-59 | 156,140 | 36,782 (23.6%) | 913,634 |
| 60-74 | 60,381 | 22,897 (37.9%) | 357,680 |
| 75-89 | 14,709 | 9,913 (67.4%) | 94,370 |
| **Sex** |  |  |  |
| Female | 240,417 | 30,295 (12.6%) | 1,377,953 |
| Male | 473,847 | 90,035 (19.0%) | 2,711,930 |
| **Race** |  |  |  |
| White | 230,953 | 40,539 (17.6%) | 1,339,033 |
| Black | 92,495 | 16,843 (18.2%) | 531,097 |
| Asian | 6,036 | 927 (15.3%) | 37,289 |
| Brown | 306,095 | 47,720 (15.6%) | 1,668,724 |
| Indigenous | 8,271 | 772 (9.3%) | 46,454 |
| Ignored | 70,414 | 13,529 (19.2%) | 467,286 |

### Mortality rate ratios forthe TB cohort

Mortality rates in the TB cohort varied by age group and years since diagnosis. **Figure 1A** reports mortality rate ratios for individuals in the study cohort compared to the general population, from the time of TB diagnosis to ten years since diagnosis. During the TB episode (first year after TB diagnosis) the MRR in the overall cohort was estimated as 11.28 (95% Confidence Interval: 11.18–11.37). This ratio dropped to 3.59 (95% CI: 3.53–3.64) in the second year after diagnosis and continued to decline monotonically, reaching 2.27 (95% CI: 2.22–2.33) in the fifth year after diagnosis and 1.46 (95% CI: 1.34–1.59) in the tenth year after diagnosis. **Figure 1B** presents MRRs estimated for each age group. For the first year since diagnosis, MRRs ranged between 4.04 (95% CI: 3.94–4.15) for individuals 75-89 years old at TB diagnosis and 24.97 (95% CI: 24.59–25.33) for individuals 30-44 years old at TB diagnosis. For each age group, MRRs declined rapidly from the first to the second year following diagnosis, then more slowly over subsequent years. In general, MRRs were highest for the 30–44 age group, and lowest for the 75–89 age group. Among the 75–89 age group, MRRs were below 1.0 (indicating lower mortality) after the 2^nd^ year following diagnosis. MRRs by year and age group are reported in **Table S2**. In addition, **Figure S1** presents MRRs by sex. In the first year following diagnosis, the MRR was higher for females (MRR: 13.59, 95% CI: 13.37–13.80) compared to males (MRR: 10.61, 95% CI: 10.51–10.72). However, this trend reversed by the third year, with males showing higher MRRs, a pattern that continued through year 10. MRRs by year and sex are reported in **Table S3**. Additionally, **Figure S2** presents MRRs by region. The South region had the highest overall MRR (MRR: 15.28, 95% CI: 14.96–15.60 in the first year), while the North region showed the lowest MRR (MRR: 9.79, 95% CI: 9.51–10.06 in the first year). MRRs by year and region are detailed in **Table S4**.

**Figure 1.**
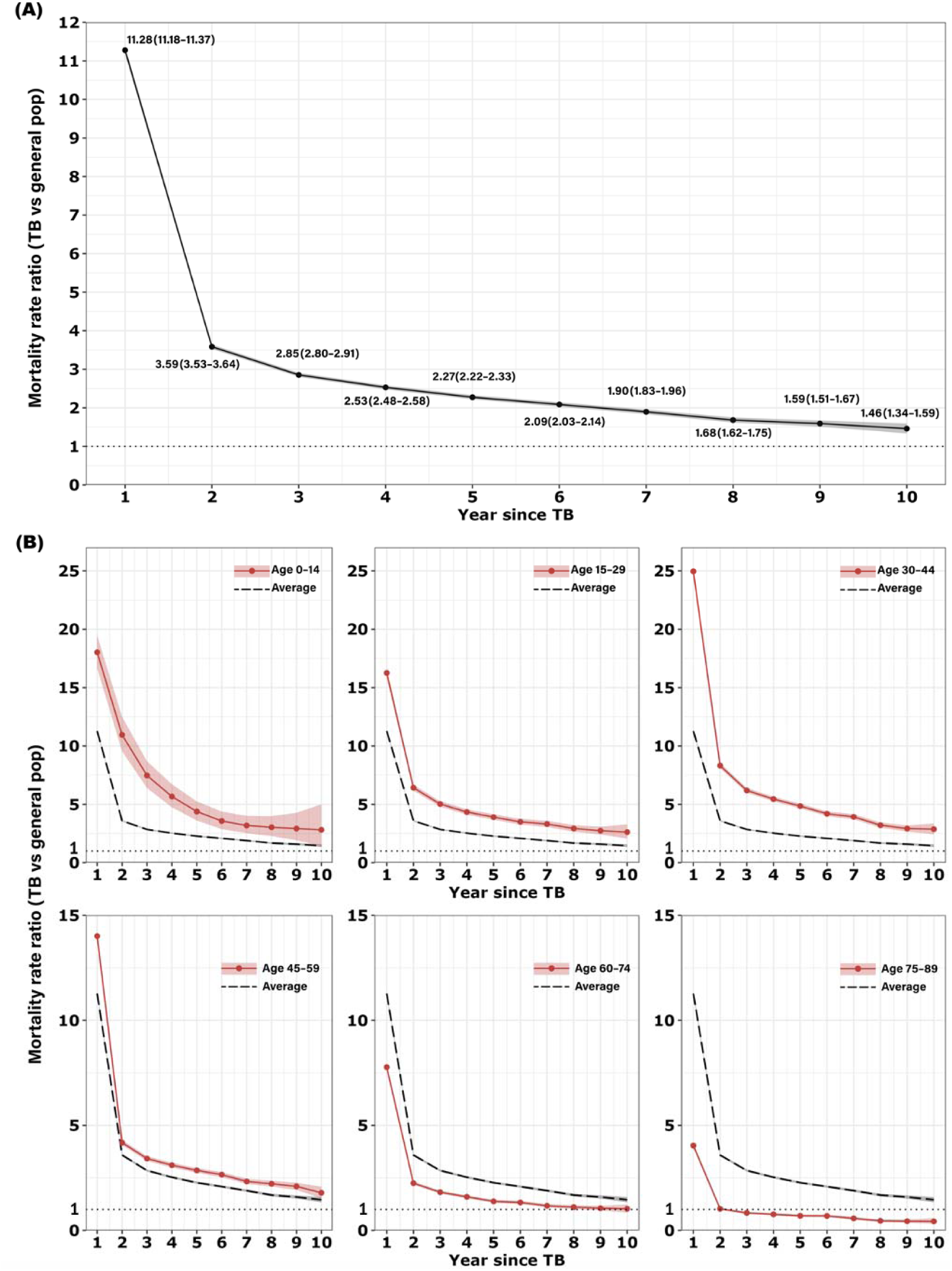
Mortality rate ratio for individuals with TB by year since TB diagnosis, for the overall TB cohort (Panel A) and by age group at TB diagnosis (Panel B), as compared to the general population.^*^ ^*^Note: In Panel B, red lines indicate the mortality rate ratio (MRR) for each age groups, and the black dashed lines show MRRs estimated for the overall population for reference. Dotted line represents MRR = 1.0 (no excess mortality

### Predictors of TB episode and post-TB mortality

The factor most strongly associated with increased mortality during the TB episode was a positive HIV diagnosis (adjusted MRR (aMRR): 5.16, 95% CI: 5.05–5.27, compared to those without HIV). Female sex (aMRR: 1.67, 95% CI: 1.64–1.70, compared to males), combined pulmonary and extrapulmonary TB (aMRR: 1.57, 95% CI: 1.52–1.62, compared to pulmonary TB), and xray evidence of other lung pathology (aMRR: 1.77, 95% CI: 1.65–1.89, compared to normal xray) were also associated with elevated mortality (**Table S5)**.

Factors associated with elevated mortality during the post-TB period (individuals surviving beyond the first 12 months after TB diagnosis) included female sex (aMRR: 1.41, 95% CI: 1.38– 1.43, compared to males), re-initiation of treatment following abandonment (aMRR: 1.78, 95% CI: 1.74–1.84, compared to new cases), positive HIV diagnosis (aMRR: 2.80, 95% CI: 2.73–2.87, compared to those without HIV), a history of alcoholism (aMRR: 1.52, 95% CI: 1.49–1.55, compared to those without such history), as well as loss to follow-up and cases with a change in regimen due to drug resistance (aMRR: 1.68, 95% CI: 1.64–1.72, and aMRR: 1.65, 95% CI: 1.53– 1.77, respectively, compared with treatment outcome of cure). **Table 2** provides details on the predictors of mortality during the post-TB period.

**Table 2.** Predictors of post-TB mortality. ^*^ Note: Post-TB period defined as >12 months after TB diagnosis. All results are adjusted for the interaction between age group and years since TB, along with race, state, and educational attainment.

|  | Univariable –<br>(95% CI) | Multivariable –<br>(95% CI) | Multivariable –<br>**without treatment**<br>**outcome** (95% CI) |
| --- | --- | --- | --- |
| **Sex** |  |  |  |
| Male | Reference | Reference | Reference |
| Female | 1.22 (1.20–1.25) | 1.41 (1.38–1.43) | 1.39 (1.37–1.42) |
| **Bacteriological test results** |  |  |  |
| Negative | Reference | Reference | Reference |
| Positive (smear or culture or Xpert) | 0.99 (0.97–1.01) | 0.99 (0.97–1.01) | 0.99 (0.97–1.02) |
| Not performed | 0.93 (0.90–0.95) | 0.97 (0.94–0.99) | 0.98 (0.95–1.01) |
| **Type of TB** |  |  |  |
| Pulmonary | Reference | Reference | Reference |
| Extrapulmonary | 0.83 (0.81–0.85) | 0.89 (0.86–0.92) | 0.87 (0.85–0.90) |
| Pulmonary + Extrapulmonary | 1.34 (1.29–1.40) | 1.04 (1.00–1.09) | 1.04 (1.00–1.09) |
| Unknown | 0.91 (0.53–1.57) | 1.25 (0.72–2.17) | 1.16 (0.67–2.00) |
| **Chest Xray** |  |  |  |
| Normal | Reference | Reference | Reference |
| Suspected | 1.20 (1.15–1.25) | 1.19 (1.14–1.24) | 1.19 (1.14–1.25) |
| Other pathology | 1.19 (1.07–1.31) | 1.20 (1.08–1.33) | 1.21 (1.09–1.34) |
| Not performed | 1.13 (1.08–1.18) | 1.11 (1.06–1.17) | 1.12 (1.07–1.17) |
| **Type of entry** |  |  |  |
| New case | Reference | Reference | Reference |
| Relapse | 1.73 (1.68–1.77) | 1.52 (1.48–1.56) | 1.58 (1.54–1.62) |
| Reentry after abandonment | 2.51 (2.44–2.57) | 1.78 (1.74–1.84) | 2.08 (2.03–2.14) |
| Unknown | 1.10 (0.93–1.30) | 1.00 (0.84–1.19) | 1.05 (0.89–1.25) |
| Transfer | 1.32 (1.27–1.37) | 1.18 (1.14–1.23) | 1.23 (1.18–1.28) |
| **Comorbid conditions** |  |  |  |
| HIV | 2.98 (2.91–3.04) | 2.80 (2.73–2.87) | 2.94 (2.87–3.02) |
| Alcoholism | 1.64 (1.60–1.67) | 1.52 (1.49–1.55) | 1.58 (1.54–1.61) |
| Diabetes | 1.25 (1.22–1.29) | 1.37 (1.33–1.41) | 1.36 (1.32–1.40) |
| Mental illness | 1.39 (1.32–1.45) | 1.20 (1.14–1.26) | 1.20 (1.14–1.26) |
| **In prison (at diagnosis)** |  |  |  |
| No | Reference | Reference | Reference |
| Yes | 1.04 (1.00–1.07) | 1.08 (1.04–1.12) | 1.05 (1.01–1.09) |
| Unknown | 0.92 (0.88–0.96) | 0.98 (0.94–1.03) | 0.98 (0.94–1.02) |
| **Recorded treatment outcome** |  |  |  |
| Cured | Reference | Reference |  |
| Loss to follow-up | 2.08 (2.04–2.12) | 1.68 (1.64–1.72) |  |
| Treatment failure | 0.79 (0.29–2.10) | 0.73 (0.27–1.94) |  |
| Transfer | 1.47 (1.43–1.52) | 1.32 (1.28–1.36) |  |
| Drug resistance + change in<br>treatment (from fixed dose<br>combination) | 2.19 (2.04–2.35) | 1.65 (1.53–1.77) |  |
| Unknown | 1.10 (1.04–1.16) | 1.06 (1.00–1.12) |  |

### Cumulative mortality

Figure 2 shows a comparison of cumulative mortality for the TB cohort and the general population over the 10-year follow-up period. In the first year, individuals in the TB cohort had a cumulative mortality of 6.82% (95% CI: 6.77 – 6.86), compared to 0.7% for the general population. Excess mortality (calculated as the difference in cumulative mortality) was 6.12% (95% CI: 6.07–6.17) at the end of the first year following diagnosis. Excess mortality grew over the post-TB period: by year five, cumulative mortality for the TB cohort reached 12.87% (95% CI: 12.73–13.01), compared to 3.69% in the general population, with 9.18% (95% CI: 9.04–9.32) excess mortality. By the end of year ten, cumulative mortality for the TB cohort was estimated as 17.88% (95% CI: 17.55–18.21), compared to 7.97% among the general population. At this point total excess mortality was 9.90% (95% CI: 9.58–10.24), with 5.22% (95% CI: 4.90–5.56) of this total resulting from post-TB mortality. Across age groups, the 45-59-year-old age group had the greatest excess mortality after 10 years (18.63%) (**Table S6 and Figure S3**). Males were estimated to have consistently higher excess mortality than females, with 11.32% (95% CI: 10.90–11.76) excess mortality after 10 years, vs. 6.95% (95% CI: 6.52–7.40) for females (**Table S7 and Figure S4**). Across regions, the South showed the highest excess mortality across all years, reaching 13.84% (95% CI: 12.89–14.83) after 10 years (**Table S8 and Figure S5**).

**Figure 2.**
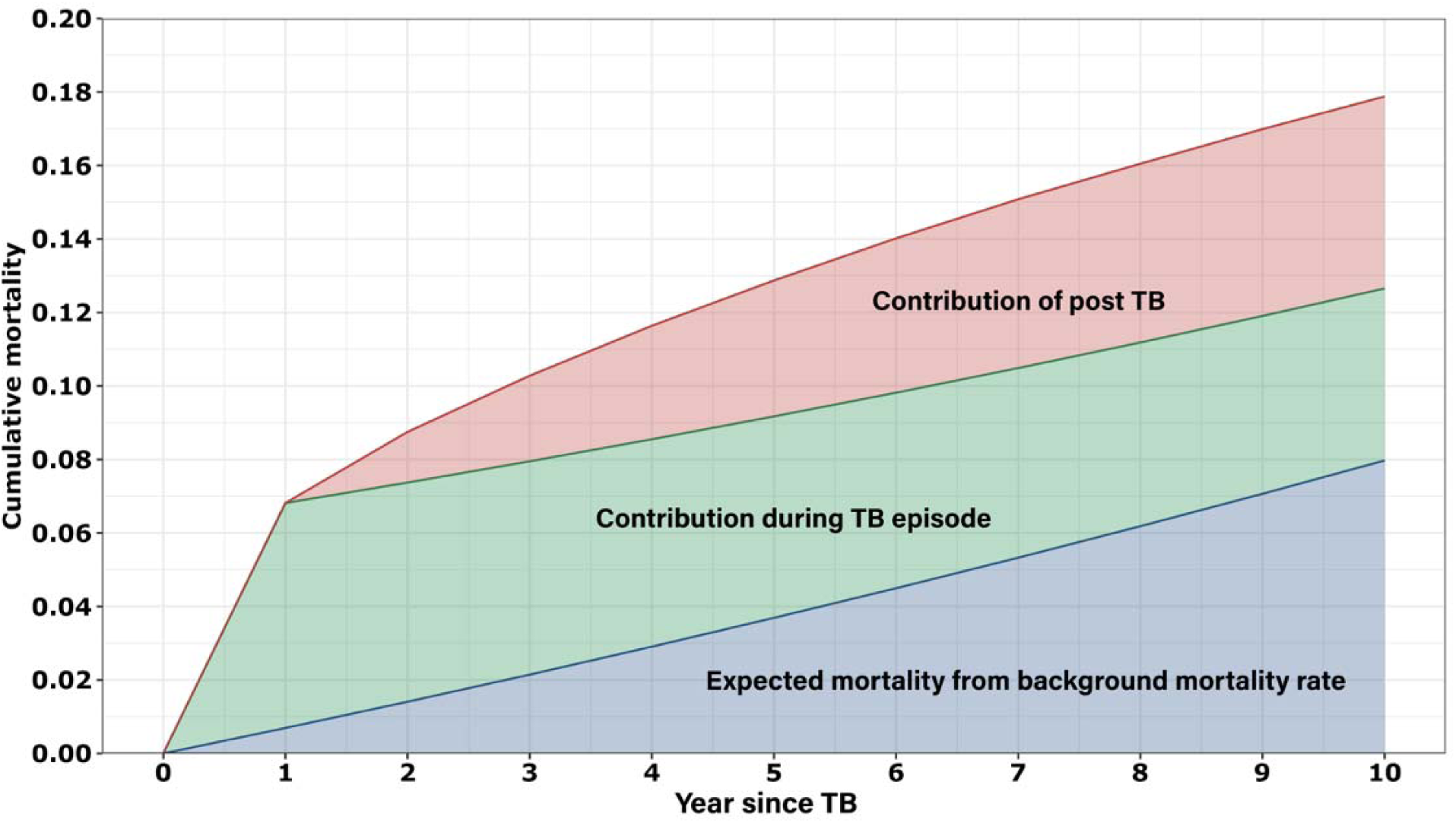
Cumulative mortality by year since TB diagnosis.^*^ ^*^Note: The blue area shows the baseline mortality rate expected in the general population without TB, the green area represents the contribution of deaths during the TB episode to overall excess mortality, and the red area indicates the contribution of mortality during the post-TB period to overall excess mortality.

### Cause of death

Reported causes of death differed markedly between the TB cohort and a matched general population sample. Figure 3 shows a comparison of the cause of death distribution between these two populations, by year since TB diagnosis. In the first year after diagnosis, TB and HIV were the leading causes of death in the TB cohort, accounting for 31.3% and 23.9% of total deaths, respectively, compared to 0.9% and 2.3% in the general population. In the tenth year after diagnosis, respiratory diseases (16.3%), cardiovascular diseases (16.1%), and cancer (13.3%) were the top causes of death for the TB cohort, compared to 9.6% for respiratory diseases, 26.3% for cardiovascular diseases, and 17.7% for cancer in the general population. **Figure S6** shows how the distribution of causes of death varied across regions.

**Figure 3.**
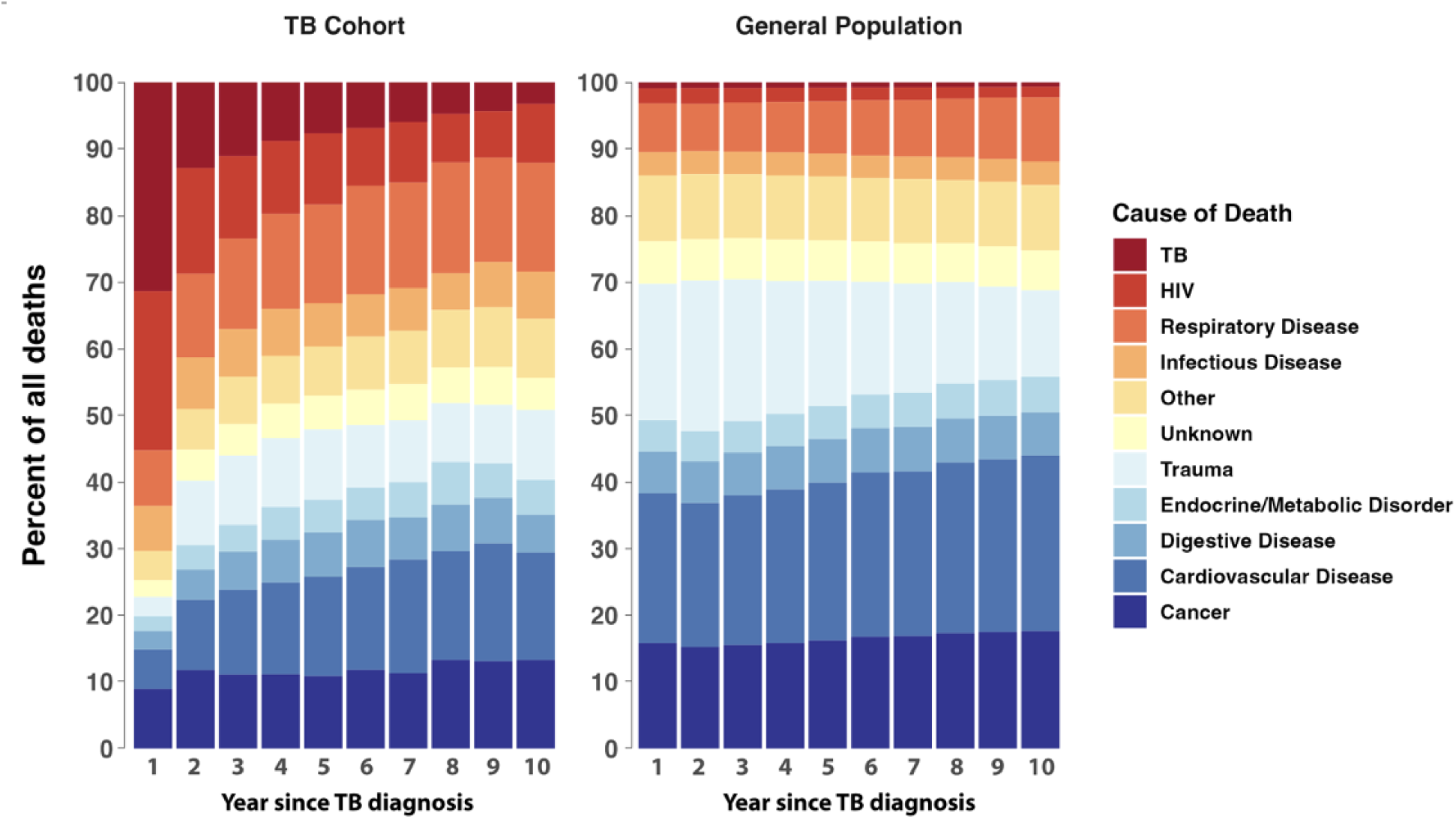
Distribution of cause of death in the TB cohort compared to a matched general population sample, by years since TB diagnosis.^*^ ^*^Note: Distribution of deaths in the general population created by weighting deaths in the national mortality registry (SIM) to match the distribution of age, sex, state of residence, and calendar year for deaths in the TB sample, for each year since diagnosis.

## DISCUSSION

In this study we assessed mortality rates among a large cohort of individuals with TB disease, up to 10 years after initial TB diagnosis. We found that individuals diagnosed with TB had significantly higher mortality compared to the general population, with mortality rates over 11 times higher in the first year after diagnosis and remaining elevated for up to 10 years after diagnosis. This extended period of elevated mortality highlights the long-term health challenges faced by TB survivors, reinforcing findings from previous studies.^5,11,12^ Notably, a recent systematic review and meta-analysis by Romanowski et al. reported a pooled standardized mortality ratio of 2.91 (95% CI: 2.21–3.84) for all-cause mortality in TB cohorts compared to the general population.^5^ While our results show similar increases in mortality for the cohort overall, they also reveal substantial variation in mortality risk during over the post-TB period, and according to patient characteristics.

MRRs estimated for each age group followed similar patterns over time, although with notable differences in magnitude, with the 30–44 age group exhibiting MRRs more than 2 times greater than the overall average. This may be due to the significant burden of comorbidities in this age group within the TB cohort, particularly the high prevalence of HIV (17.9%) and alcoholism (19.9%), both of which are strong predictors of mortality. In contrast, the 75–89 age group had the lowest MRRs, which could be related to elevated baseline mortality rates in the older population, as well as survivorship bias.^19^

This study also identified key predictors of long-term mortality among individuals who survived the first 12 months after TB diagnosis (post-TB). Relapse, loss to follow-up, and drug-resistant TB were each strongly associated with increased mortality within the TB cohort, reflecting the harmful effects of interrupted or unsuccessful TB treatment, associated with ongoing disease progression and other complications.^20–22^ In addition, HIV infection was one of the strongest predictors of mortality, with HIV-positive individuals having a post-TB mortality rate 2.83 times that of HIV-negative individuals. This finding reinforces the well-established interaction between TB and HIV, where co-infection exacerbates the severity and progression of both diseases, leading to higher mortality.^23–25^ In addition to HIV, individuals with a history of alcoholism had a higher risk of death. This could be due to the harmful effects of alcohol on the immune system and its potential to worsen TB outcomes through poor treatment adherence and increased risk of complications.^26,27^ We also found MRRs to be higher for females compared to males. This result is surprising, given other study results finding females to have lower mortality during TB treatment,^28,29^ and the difference remained after controlling for a range of other patient covariates. Additionally, the highest MRRs were observed in the South region, corresponding to elevated TB–HIV coinfection rates, which reach 15.2%—one of the highest in the country. ^30,31^

The results for cumulative mortality highlight both the immediate and long-term excess mortality burden among TB survivors. While there was substantial excess mortality in the TB cohort during the first 12 months following diagnosis, mortality during the post-TB period represented 53% of overall excess mortality. This finding aligns with recent global estimates of the lifetime burden of TB disease, which attributed 47% of the total TB-related disease burden in 2019 to post-TB sequelae.^12^ These results highlight the lasting effects of TB on long-term health, even after the TB episode.

Lastly, the cause of death analysis highlights the evolving health risks for TB survivors over time. In the first year after diagnosis, TB and HIV were leading causes of death. By the tenth year, however, the main causes of death among TB survivors had shifted to respiratory diseases, cardiovascular diseases, and cancer. This is consistent with recent findings indicating that a substantial portion (20%) of post-treatment deaths are attributable to cardiovascular disease.^5^ This transition in causes of death among the TB cohort—from TB and HIV in the early years to chronic conditions in later years—reflects the complex health challenges TB survivors face as they age. These findings reflects not only the immediate risks posed by TB but also the long-term consequences of TB sequelae, which increase survivors’ susceptibility to other diseases, including non-communicable diseases.^32^ This also underscores the importance of ongoing healthcare support and preventive strategies that extend well beyond the completion of TB treatment to manage the chronic health challenges TB survivors encounter over time.^33^

This study has several limitations. First, while the dataset used is large and nationally representative, our results rely on the accuracy of the SINAN and SIM data. Any misclassification of TB status or cause of death in the data could introduce bias into our estimates. Second, while we defined the ’post-TB period’ as beginning 12 months following diagnosis, individuals being treated for drug-resistant TB may receive regimens of 18-24 months in length. We chose 12 months to have a consistent definition for all patients, and since patients receiving long regimens represent a small minority of the treatment cohort. Third, we were unable to account for individuals who may have emigrated from Brazil during the study period. However, based on contemporary estimates from the International Organization for Migration (IOM), the annual emigration rate in Brazil was approximately 0.8% as of 2019.^34^ Given this low rate, it is unlikely that emigration had a major impact on analytic results. Fourth, we controlled for state using the residence at the time of diagnosis. Although there was a low discrepancy (1.3%) between the state of residence at diagnosis and at death, this may not fully account for the living conditions the TB cohort experienced until death, which could impact the mortality rate ratio calculations or comparisons of cause of death distributions. Finally, while our study adjusted for age, sex, state, and year of death to calculate the MRRs of the TB cohort compared to the general population’s background mortality rates, we were unable to match on other socioeconomic or demographic variables, such as income or occupation. These unmeasured factors may have contributed to the mortality differences observed between the groups, indicating that our results reflect not only the causal effect of TB, but the combined effect of TB and these underlying socioeconomic determinants.

Despite these limitations, this study represents one of the most comprehensive analyses of long-term survival among individuals who develop TB disease, and it is among the first to explore how mortality risk ratios (MRRs) of TB survivors vary by years since diagnosis compared to the general population. Drawing on data from over 800,000 individuals and more than 4 million years of follow-up, we were able to examine excess mortality across a wide range of patient characteristics—including age, sex, and clinical and demographic factors—at a level of detail not possible in smaller studies. The findings from our study highlight the ongoing healthcare needs of TB survivors, a challenge not unique to Brazil but relevant to many other countries facing significant TB burdens.^10^

## Data Availability

All data produced in the present study are available upon reasonable request to the authors.

## SUPPLEMENT

**Table S1.** Prevalence of comorbid conditions in the TB cohort by age group.

|  | HIV | Diabetes | Alcoholism | Mental illness |
| --- | --- | --- | --- | --- |
| **Overall (%)** | 11.00 | 6.21 | 15.52 | 2.26 |
| **Age group** |  |  |  |  |
| 0-14 | 4.58 | 0.95 | 1.72 | 1.22 |
| 15-29 | 8.06 | 1.07 | 8.38 | 1.64 |
| 30-44 | 17.93 | 3.70 | 19.94 | 2.55 |
| 45-59 | 10.67 | 11.53 | 22.72 | 2.89 |
| 60-74 | 3.42 | 16.51 | 13.20 | 2.10 |
| 75-89 | 1.17 | 13.05 | 4.78 | 1.75 |

**Table S2.** Mortality rate ratios by year since diagnosis and age group.

|  | Year 1<br>MRR (95% CI) | Year 2<br>MRR (95% CI) | Year 5<br>MRR (95% CI) | Year 10<br>MRR (95% CI) |
| --- | --- | --- | --- | --- |
| **Overall** | 11.28 (11.18–11.37) | 3.58 (3.53–3.64) | 2.27 (2.22–2.33) | 1.46 (1.34–1.59) |
| **Age group** |  |  |  |  |
| 0-14 | 18.20 (16.84–19.65) | 10.36 (8.53–12.42) | 4.22 (3.00–5.76) | 1.95 (0.39–5.92) |
| 15-29 | 16.28 (15.90–16.66) | 6.37 (6.11–6.63) | 3.92 (3.67–4.18) | 2.63 (2.04–3.34) |
| 30-44 | 24.97 (24.59–25.33) | 8.29 (8.06–8.52) | 4.86 (4.65–5.07) | 2.87 (2.44–3.38) |
| 45-59 | 14.02 (13.81–14.22) | 4.16 (4.04–4.28) | 2.86 (2.74–2.97) | 1.78 (1.52–2.08) |
| 60-74 | 7.77 (7.64–7.91) | 2.24 (2.16–2.32) | 1.38 (1.31–1.46) | 1.03 (0.85–1.25) |
| 75-89 | 4.04 (3.94–4.15) | 1.02 (0.96–1.08) | 0.70 (0.63–0.75) | 0.43 (0.30–0.59) |

MRR = mortality rate ratio. CI = confidence interval.

**Table S3.** Mortality rate ratios by year since diagnosis and sex.

|  | Year 1<br>MRR (95% CI) | Year 2<br>MRR (95% CI) | Year 5<br>MRR (95% CI) | Year 10<br>MRR (95% CI) |
| --- | --- | --- | --- | --- |
| **Overall** | 11.28 (11.18–11.37) | 3.58 (3.53–3.64) | 2.27 (2.22–2.33) | 1.46 (1.34–1.59) |
| **Sex** |  |  |  |  |
| Female | 13.59 (13.37–13.80) | 3.97 (3.85–4.10) | 2.15 (2.05–2.26) | 1.30 (1.09–1.55) |
| Male | 10.61 (10.51–10.72) | 3.48 (3.42–3.54) | 2.31 (2.25–2.37) | 1.52 (1.37–1.67) |

MRR = mortality rate ratio. CI = confidence interval.

**Table S4.** Mortality rate ratios by year and region.

|  | Year 1<br>MRR (95% CI) | Year 2<br>MRR (95% CI) | Year 5<br>MRR (95% CI) | Year 10<br>MRR (95% CI) |
| --- | --- | --- | --- | --- |
| **Overall** | 11.28 (11.18–11.37) | 3.58 (3.53–3.64) | 2.27 (2.22–2.33) | 1.46 (1.34–1.59) |
| **Region** |  |  |  |  |
| North | 9.79 (9.51–10.06) | 3.00 (2.83–3.16) | 1.73 (1.59–1.89) | 1.10 (0.79–1.50) |
| Northeast | 9.88 (9.72–10.04) | 3.26 (3.16–3.36) | 2.02 (1.93–2.11) | 1.13 (0.93–1.35) |
| Central West | 10.77 (10.38–11.17) | 3.45 (3.22–3.70) | 2.24 (2.01–2.49) | 1.00 (0.60–1.59) |
| Southeast | 11.48 (11.34–11.62) | 3.56 (3.47–3.64) | 2.36 (2.28–2.44) | 1.74 (1.54–1.96) |
| South | 15.28 (14.96–15.60) | 5.01 (4.83–5.21) | 3.06 (2.88–3.25) | 1.82 (1.43–2.29) |

MRR = mortality rate ratio. CI = confidence interval.

**Table S5.**
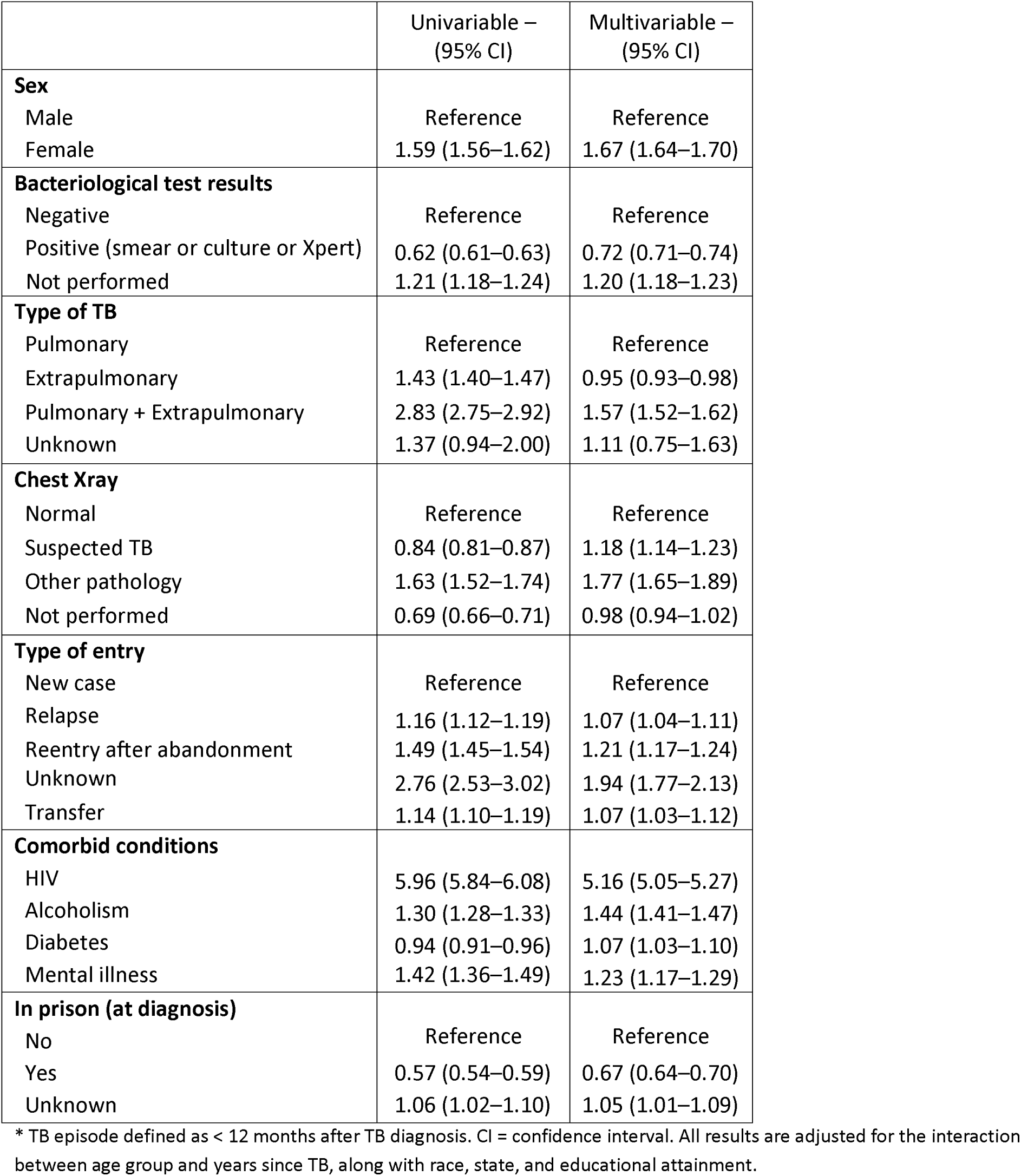
Mortality rate ratios during the TB episode for patient characteristics.*

**Table S6.**
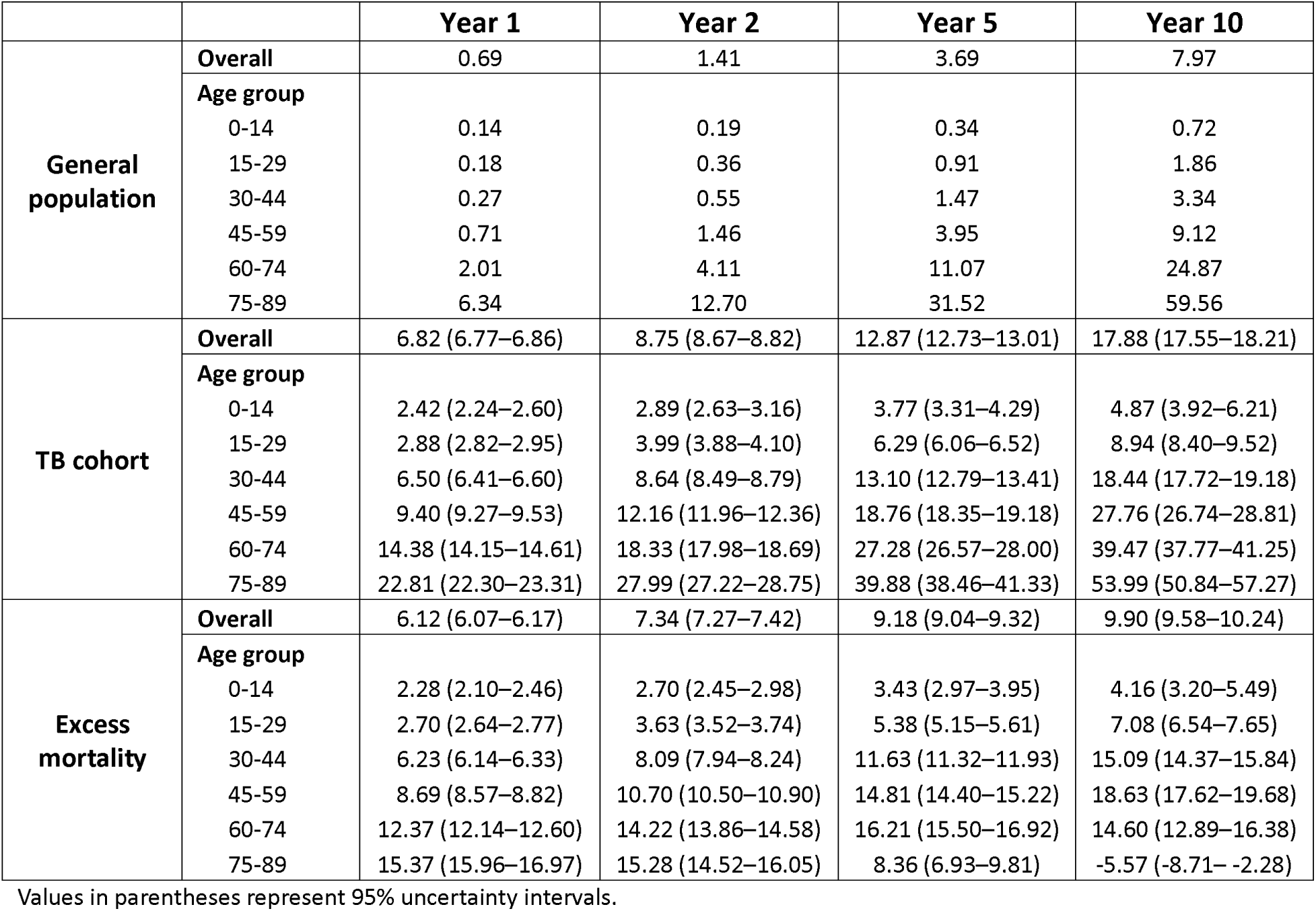
Cumulative mortality (%) by year since diagnosis and age group.

|  |  | **Year 1** | **Year 2** | **Year 5** | **Year 10** |
| --- | --- | --- | --- | --- | --- |
| **General population** | **Overall** | 0.69 | 1.41 | 3.69 | 7.97 |
|  | **Age group** |  |  |  |  |
|  | 0-14 | 0.14 | 0.19 | 0.34 | 0.72 |
|  | 15-29 | 0.18 | 0.36 | 0.91 | 1.86 |
|  | 30-44 | 0.27 | 0.55 | 1.47 | 3.34 |
|  | 45-59 | 0.71 | 1.46 | 3.95 | 9.12 |
|  | 60-74 | 2.01 | 4.11 | 11.07 | 24.87 |
|  | 75-89 | 6.34 | 12.70 | 31.52 | 59.56 |
| **TB cohort** | **Overall** | 6.82 (6.77–6.86) | 8.75 (8.67–8.82) | 12.87 (12.73–13.01) | 17.88 (17.55–18.21) |
|  | **Age group** |  |  |  |  |
|  | 0-14 | 2.42 (2.24–2.60) | 2.89 (2.63–3.16) | 3.77 (3.31–4.29) | 4.87 (3.92–6.21) |
|  | 15-29 | 2.88 (2.82–2.95) | 3.99 (3.88–4.10) | 6.29 (6.06–6.52) | 8.94 (8.40–9.52) |
|  | 30-44 | 6.50 (6.41–6.60) | 8.64 (8.49–8.79) | 13.10 (12.79–13.41) | 18.44 (17.72–19.18) |
|  | 45-59 | 9.40 (9.27–9.53) | 12.16 (11.96–12.36) | 18.76 (18.35–19.18) | 27.76 (26.74–28.81) |
|  | 60-74 | 14.38 (14.15–14.61) | 18.33 (17.98–18.69) | 27.28 (26.57–28.00) | 39.47 (37.77–41.25) |
|  | 75-89 | 22.81 (22.30–23.31) | 27.99 (27.22–28.75) | 39.88 (38.46–41.33) | 53.99 (50.84–57.27) |
| **Excess mortality** | **Overall** | 6.12 (6.07–6.17) | 7.34 (7.27–7.42) | 9.18 (9.04–9.32) | 9.90 (9.58–10.24) |
|  | **Age group** |  |  |  |  |
|  | 0-14 | 2.28 (2.10–2.46) | 2.70 (2.45–2.98) | 3.43 (2.97–3.95) | 4.16 (3.20–5.49) |
|  | 15-29 | 2.70 (2.64–2.77) | 3.63 (3.52–3.74) | 5.38 (5.15–5.61) | 7.08 (6.54–7.65) |
|  | 30-44 | 6.23 (6.14–6.33) | 8.09 (7.94–8.24) | 11.63 (11.32–11.93) | 15.09 (14.37–15.84) |
|  | 45-59 | 8.69 (8.57–8.82) | 10.70 (10.50–10.90) | 14.81 (14.40–15.22) | 18.63 (17.62–19.68) |
|  | 60-74 | 12.37 (12.14–12.60) | 14.22 (13.86–14.58) | 16.21 (15.50–16.92) | 14.60 (12.89–16.38) |
|  | 75-89 | 15.37 (15.96–16.97) | 15.28 (14.52–16.05) | 8.36 (6.93–9.81) | -5.57 (-8.71– -2.28) |

Values in parentheses represent 95% uncertainty intervals.

**Table S7.** Cumulative and excess mortality (%) by year since diagnosis and sex.

|  |  | **Year 1** | **Year 2** | **Year 5** | **Year 10** |
| --- | --- | --- | --- | --- | --- |
| **General population** | **Overall** | 0.69 | 1.41 | 3.69 | 7.97 |
|  | Female | 0.47 | 0.97 | 2.59 | 5.76 |
|  | Male | 0.80 | 1.62 | 4.22 | 9.04 |
| **TB cohort** | **Overall** | 6.82 (6.77–6.86) | 8.75 (8.67–8.82) | 12.87 (12.73–13.01) | 17.88 (17.55–18.21) |
|  | Female | 5.43 (5.35–5.50) | 6.82 (6.71–6.93) | 9.50 (9.30–9.70) | 12.71 (12.28–13.15) |
|  | Male | 7.46 (7.40–7.52) | 9.65 (9.56–9.75) | 14.47 (14.28–14.66) | 20.36 (19.93–20.79) |
| **Excess mortality** | **Overall** | 6.12 (6.07–6.17) | 7.34 (7.27–7.42) | 9.18 (9.04–9.32) | 9.90 (9.58–10.24) |
|  | Female | 4.95 (4.88–5.03) | 5.85 (5.74–5.96) | 6.91 (6.72–7.11) | 6.95 (6.52–7.40) |
|  | Male | 6.66 (6.60–6.72) | 8.03 (7.94–8.13) | 10.25 (10.06–10.43) | 11.32 (10.90–11.76) |

Values in parentheses represent 95% uncertainty intervals.

**Table S8.** Cumulative mortality (%) by year since diagnosis and region.

|  |  | **Year 1** | **Year 2** | **Year 5** | **Year 10** |
| --- | --- | --- | --- | --- | --- |
| **General population** | **Overall** | 0.69 | 1.41 | 3.69 | 7.97 |
|  | **Region** |  |  |  |  |
|  | North | 0.67 | 1.36 | 3.57 | 7.72 |
|  | Northeast | 0.74 | 1.51 | 3.93 | 8.47 |
|  | Central-West | 0.80 | 1.62 | 4.21 | 9.04 |
|  | Southeast | 0.67 | 1.35 | 3.55 | 7.69 |
|  | South | 0.67 | 1.36 | 3.57 | 7.78 |
| **TB cohort** | **Overall** | 6.82 (6.77–6.86) | 8.75 (8.67–8.82) | 12.87 (12.73–13.01) | 17.88 (17.55–18.21) |
|  | **Region** |  |  |  |  |
|  | North | 5.79 (5.64–5.93) | 7.39 (7.17–7.60) | 10.58 (10.17–11.00) | 14.34 (13.42–15.33) |
|  | Northeast | 6.43 (6.34–6.52) | 8.33 (8.19–8.47) | 12.36 (12.10–12.63) | 17.09 (16.49–17.71) |
|  | Central-West | 7.41 (7.17–7.64) | 9.50 (9.14–9.86) | 13.80 (13.13–14.48) | 18.56 (17.10–20.16) |
|  | Southeast | 6.70 (6.63–6.78) | 8.57 (8.46–8.69) | 12.74 (12.53–12.96) | 18.08 (17.59–18.59) |
|  | South | 8.62 (8.47–8.77) | 11.07 (10.85–11.30) | 15.96 (15.53–16.39) | 21.61 (20.67–22.60) |
| **Excess mortality** | **Overall** | 6.12 (6.07–6.17) | 7.34 (7.27–7.42) | 9.18 (9.04–9.32) | 9.90 (9.58–10.24) |
|  | **Region** |  |  |  |  |
|  | North | 5.11 (4.97–5.26) | 6.02 (5.81–6.24) | 7.00 (6.60–7.42) | 6.62 (5.69–7.61) |
|  | Northeast | 5.69 (5.60–5.78) | 6.83 (6.69–6.97) | 8.43 (8.16–8.70) | 8.62 (8.02–9.24) |
|  | Central-West | 6.61 (6.38–6.85) | 7.88 (7.53–8.24) | 9.59 (8.92–10.27) | 9.52 (8.06–11.12) |
|  | Southeast | 6.04 (5.97–6.11) | 7.22 (7.11–7.33) | 9.19 (8.98–9.40) | 10.39 (9.90–10.90) |
|  | South | 7.95 (7.80–8.10) | 9.71 (9.49–9.94) | 12.38 (11.96–12.81) | 13.84 (12.89–14.83) |

Values in parentheses represent 95% uncertainty intervals.

**Figure S1.**
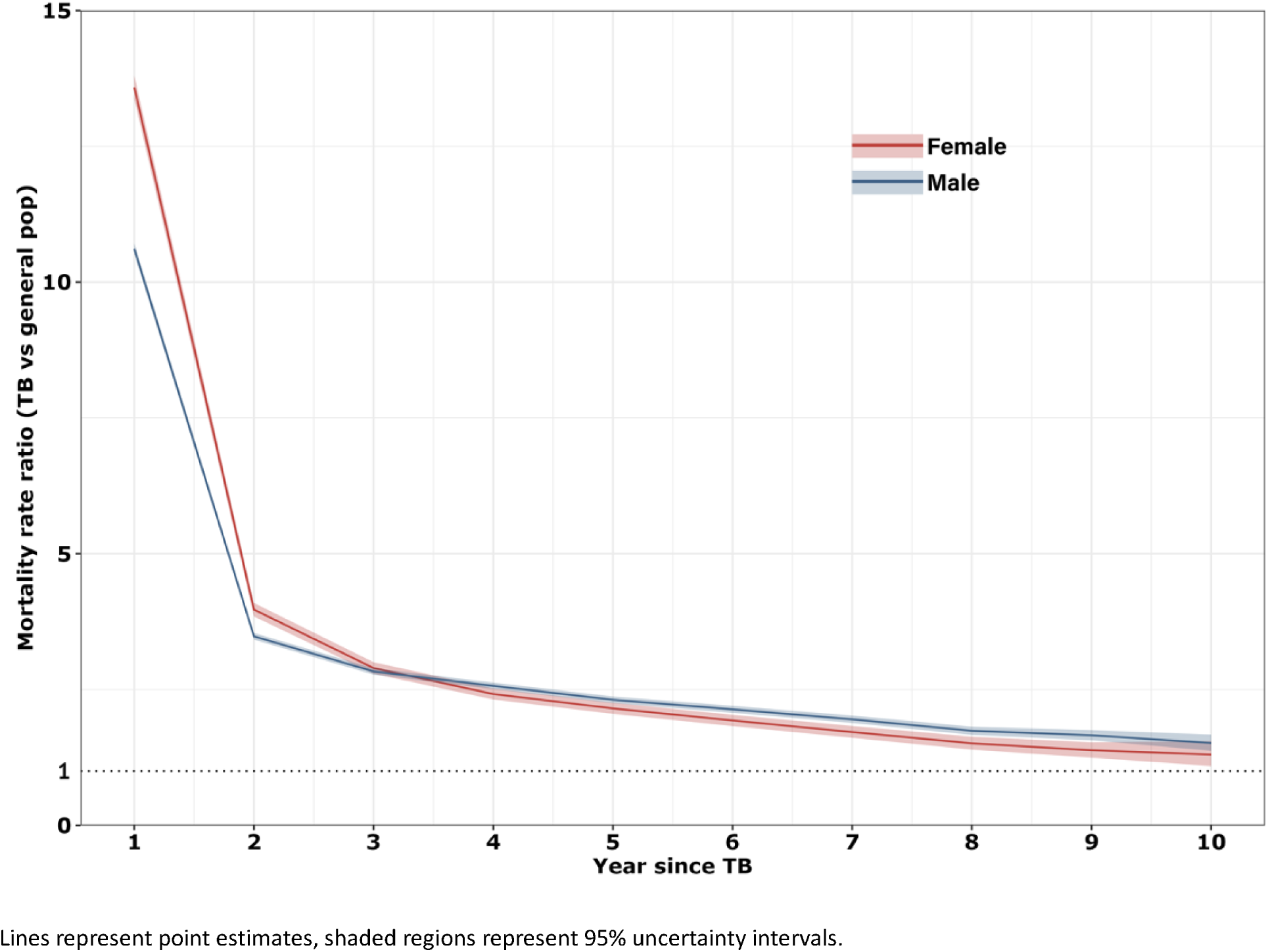
Mortality rate ratios for individuals with TB by year since diagnosis and sex, as compared to the general population.

**Figure S2.**
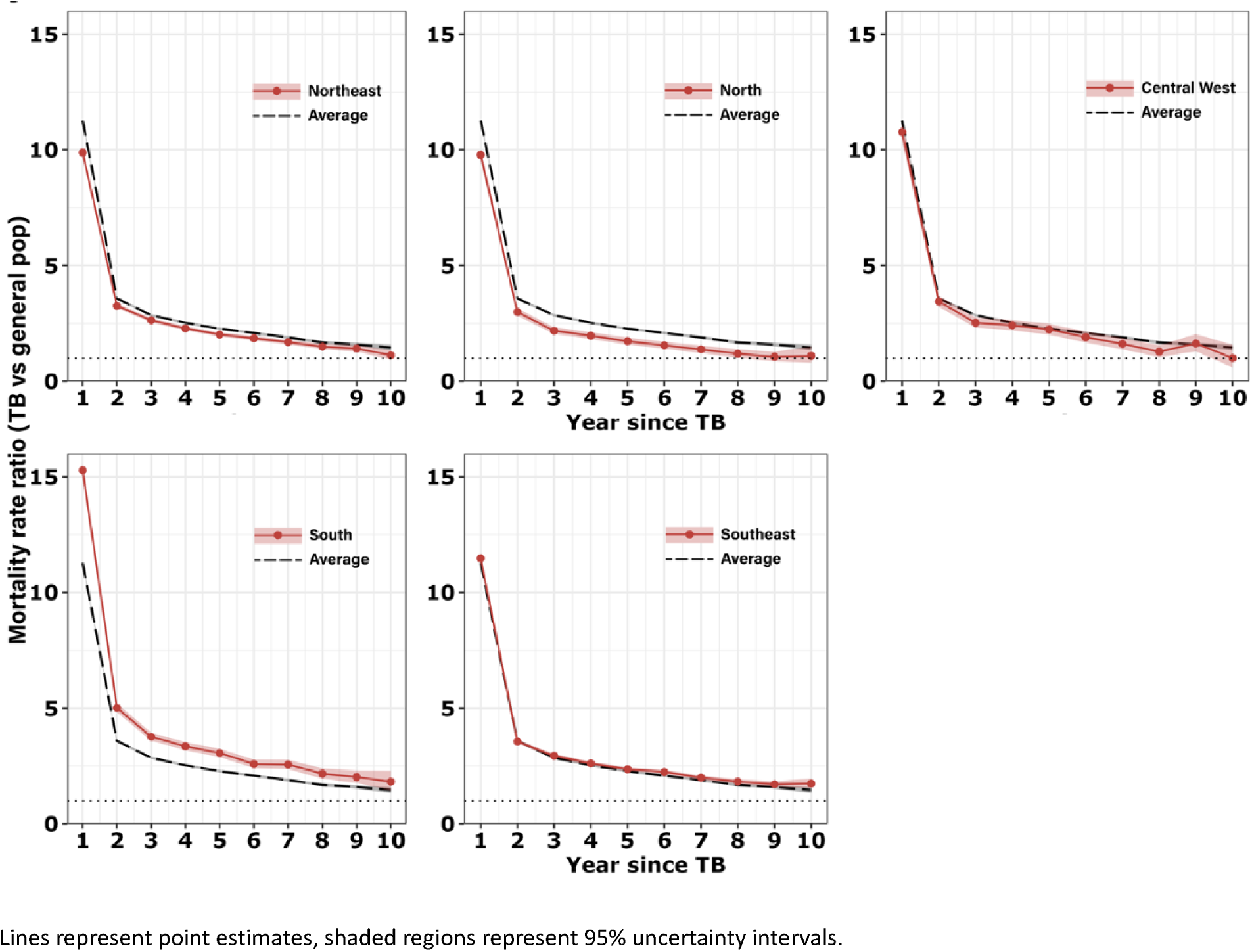
Mortality rate ratios for individuals with TB by year since diagnosis and by region, as compared to the general population.

**Figure S3.**
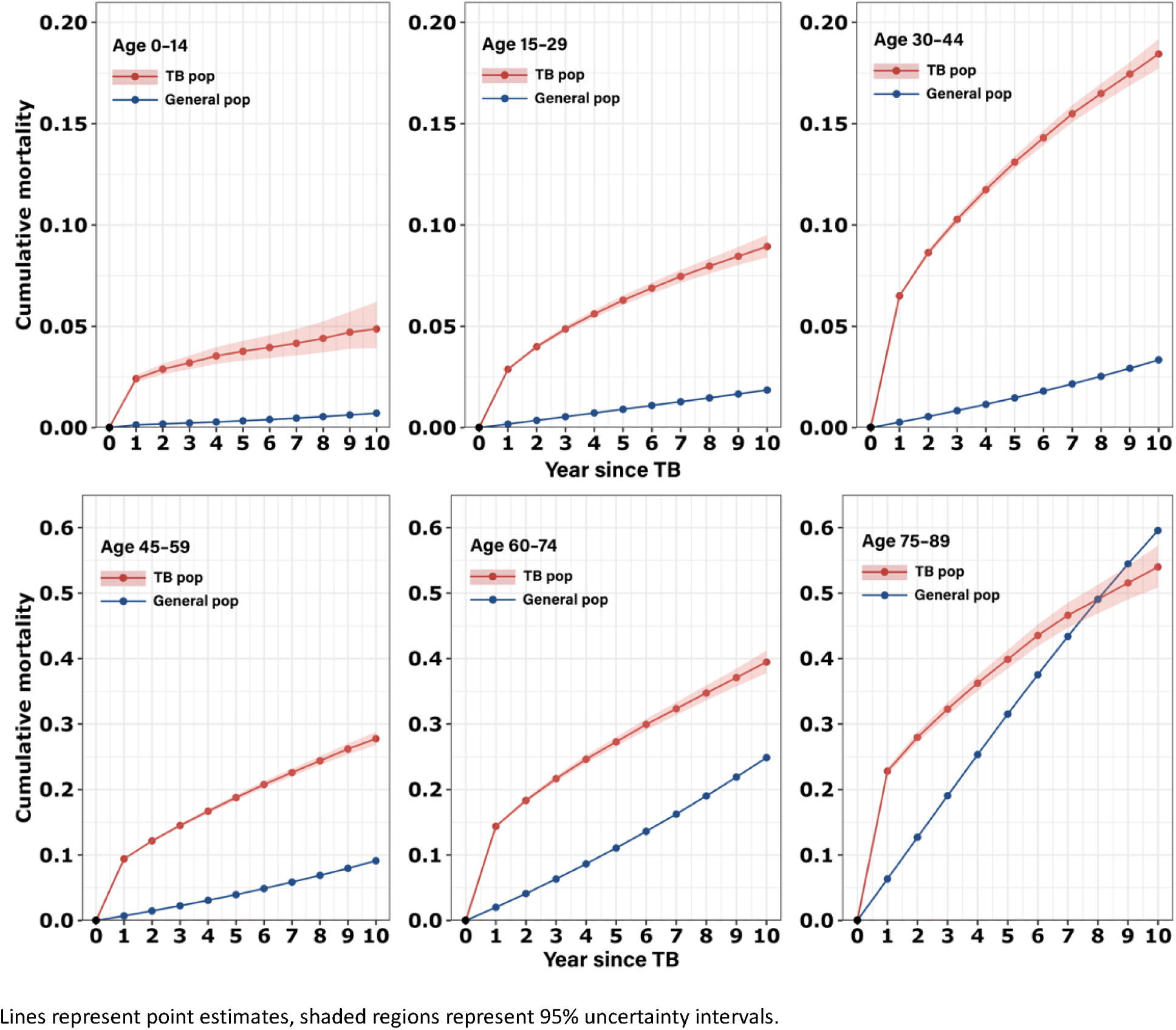
Cumulative mortality by age group.

**Figure S4.**
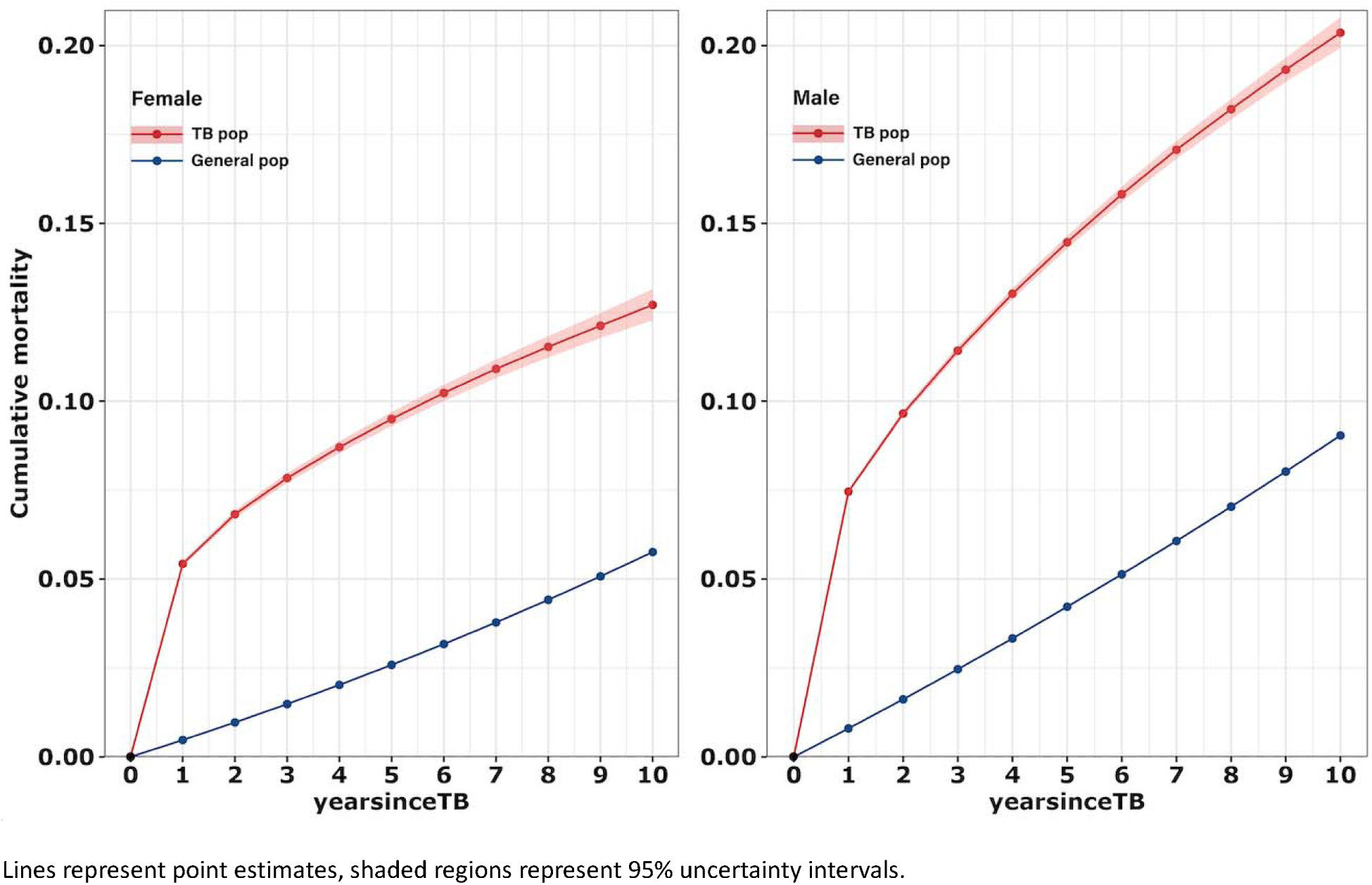
Cumulative mortality by sex.

**Figure S5.**
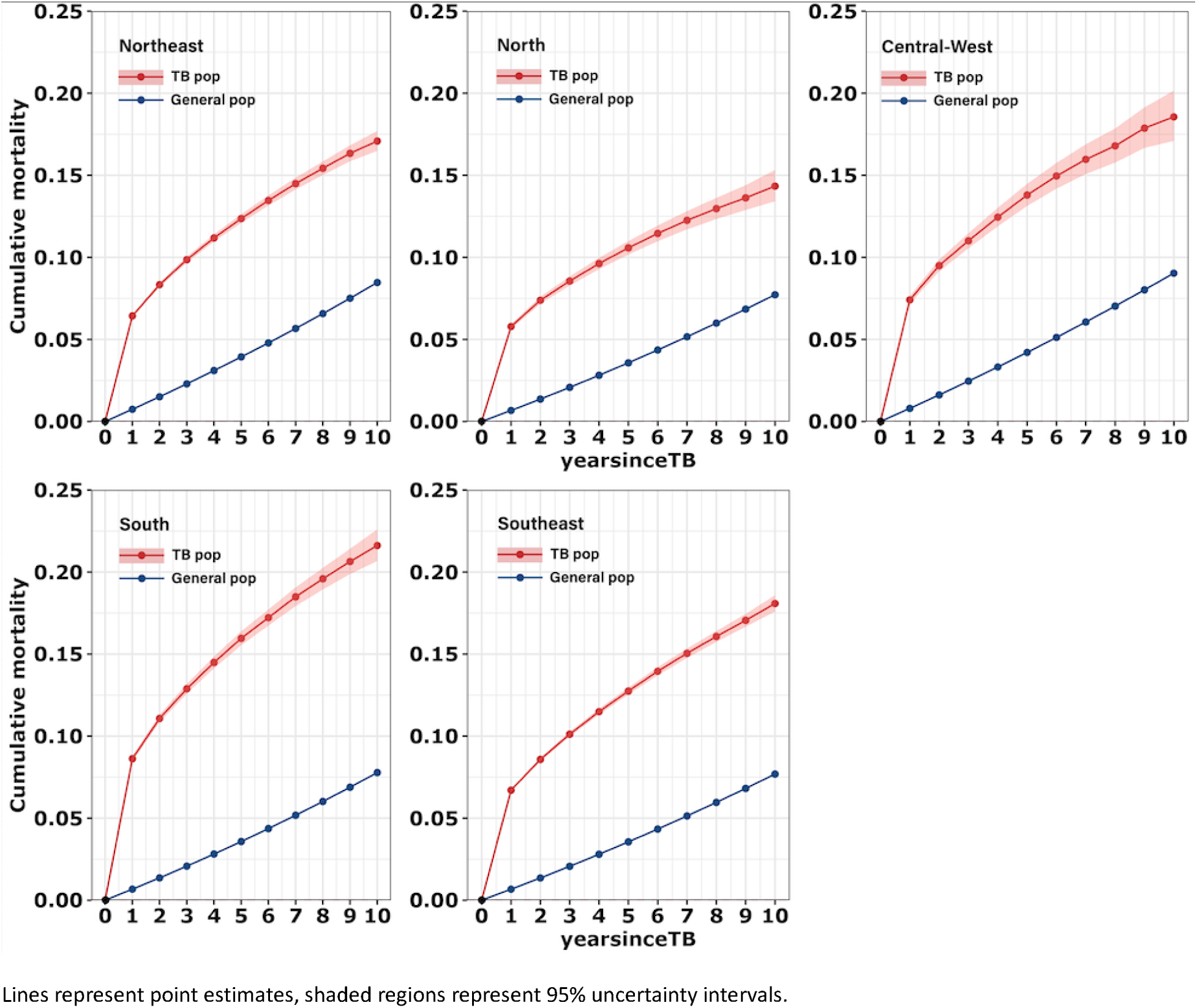
Cumulative mortality by region.

**Figure S6.**
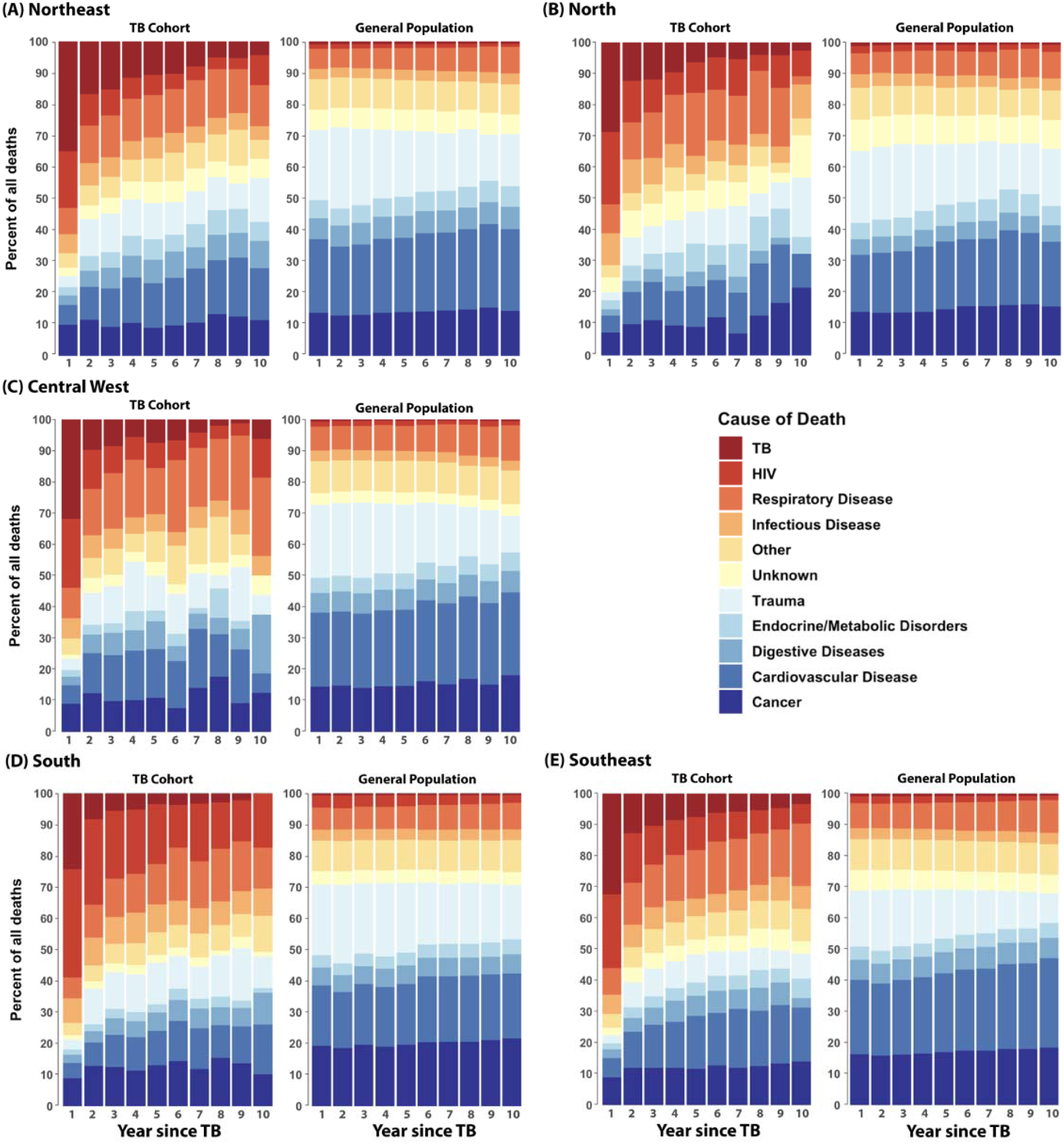
Distribution of cause of death in the TB cohort compared to a matched general population sample, by years since TB diagnosis, by region.

